# Maternal plasma lipids are involved in the pathogenesis of preterm birth

**DOI:** 10.1101/2021.07.04.21259965

**Authors:** Yile Chen, Bing He, Yu Liu, Max T. Aung, Zaira Rosario-Pabón, Carmen M. Vélez-Vega, Akram Alshawabkeh, José F. Cordero, John D. Meeker, Lana X. Garmire

**Affiliations:** Deparment of Computational Medicine and Bioinformatics, University of Michigan, Ann Arbor, 48105, USA; Program on Reproductive Health and the Environment, Department of Obstetrics, Gynecology, and Reproductive Sciences, University of California, San Francisco, School of Medicine, San Francisco, CA, USA; University of Puerto Rico Graduate School of Public Health, UPR Medical Sciences Campus, San Juan, Puerto Rico; College of Engineering, Northeastern University, Boston, Massachusetts, United States; Department of Epidemiology and Biostatistics, University of Georgia, Athens, Georgia, United States; Deparment of Environmental and Health Sciences, School of Public Health, University of Michigan, Ann Arbor, 48109, USA

**Author notes:** These authors contributed equally to the work.

**Keywords:** preterm, metabolomics, lipid, metabolic pathway, biomarkers, network, fatty acid

## Abstract

**Background:** Preterm birth is defined by the onset of labor at a gestational age shorter than 37 weeks and it can lead to premature birth and impose a threat to newborns’ health. The Puerto Rico PROTECT cohort is a well-characterized prospective birth cohort that was designed to investigate environmental and social contributors to preterm birth in Puerto Rico, where preterm birth rates have been elevated in recent decades. To elucidate possible relationships between metabolites and preterm birth in this cohort, we conducted a nested case-control study to conduct untargeted metabolomic characterization of maternal plasma of 31 preterm birth women and 69 full-term labor controls at 24-28 gestational weeks.

**Results:** A total of 333 metabolites were identified and annotated with liquid chromatography/mass spectrometry. Subsequent weighted gene correlation network analysis shows the fatty acid and carene enriched module has a significant positive association (p-value=8e-04) with preterm birth. After controlling for potential clinical confounders, a total of 38 metabolites demonstrated significant changes uniquely associated with preterm birth, where 17 of them were preterm biomarkers. Among seven machine-learning classifiers, application of random forest achieved the highly accurate and specific prediction (AUC = 0.92) for preterm birth in testing data, demonstrating their strong potential as biomarkers for preterm births. The 17 preterm biomarkers are involved in cell signaling, lipid metabolism, and lipid peroxidation functions. Further causality analysis infers that suberic acid upregulates several fatty acids to promote preterm birth.

**Conclusions:** Altogether, this study demonstrates the involvement of lipids, particularly fatty acids, in the pathogenesis of preterm birth.

## Introduction

Preterm birth is defined as deliveries that occur prior to 37 weeks of gestation, and it is one of the leading causes of newborn mortality and morbidity [1]. We previously reported that the rates of preterm birth in Puerto Rico are among the highest observed worldwide, reaching 18% [2]. The Puerto Rico PROTECT cohort, herein referred to as the PROTECT cohort, was established to study the etiology of preterm birth and the risk factors associated with it. Factors such as higher maternal age [3], smoking history [4] and lower socioeconomic status, particularly as indicated by education level and income level [5] have been reported to be associated with adverse labor outcome[2]. Additionally, we conducted an environmental exposure study in PROTECT and found that phthalate metabolites were positively associated with preterm birth [6]. Endogenous metabolites derived from important biological processes (e.g. lipolysis, glycolysis) may provide critical insight into the etiology of antecedent mechanisms of preterm birth [7], therefore, we conducted a metabolomics study within the PROTECT cohort to establish a potential link between metabolites and preterm birth.

Metabolomics provides compositional and quantitative information about the state of an organism or cell at the macromolecular level [8]. Blood metabolomics has been used to identify biomarkers and potential molecular mechanisms for various diseases and conditions, such as aging [9], acute-on-chronic liver failure [10], hypertension and blood pressure progression [11]. Biomarkers of preterm birth have been discovered in the amniotic fluid, maternal urine/maternal blood, cervicovaginal fluid [7]. Decreased phosphocholine (PC) [12] and increased levels of acylglycerophosphoserines (PS), diacylglycerophosphoethanolamines (PE), phosphatidyinositol (PI), and phosphatiduglecerol (PG) were observed in maternal blood samples from women with preterm birth [13]. In a previous lipidomic analysis in the PROTECT cohort, we have also observed signals between maternal free fatty acids and phospholipids (plasmenyl-phosphatidylethanolamines) and spontaneous preterm birth [14]. We sought to expand on this body of evidence and explore greater coverage of metabolic pathways and conducted this study to explore the potential roles of lipids in the preterm birth.

The samples utilized in this study were maternal plasma collected in gestational weeks 24-28 from the women, who went on to experience preterm birth (N = 31) or full-term healthy deliveries (N = 69). Untargeted metabolomics LC-MS/MS assays were performed on these samples, followed by bioinformatics analysis. Our goals are the following: (1) identify metabolites and metabolomic pathways that are associated with preterm birth; (2) elucidate metabolomic processes that may have a causal relationship with preterm birth; (3) seek early gestational metabolomic biomarkers (week 24-28) that are predictive of preterm birth.

## Materials and Methods

### Study population

This study was conducted in an exploratory sample of the PROTECT cohort, which obtained its own institutional review boards (IRB) approval. At the time of this study, the parent cohort consisted of 812 pregnant women, from which we randomly sampled 31 women who experienced preterm birth and 69 full-term controls for metabolomic analysis. Recruitment of the PROTECT cohort is ongoing and began in 2010. It’s funded by the National Institute of Environmental Health Sciences Superfund Research Program. Participants were recruited in the first or second trimester of pregnancy (median 14 weeks gestation). Inclusion criteria for recruitment were: being 18-40 years of age; having residence in the Northern Karst aquifer region; disuse of oral contraceptives three months before pregnancy; disuse of *in vitro* fertilization; and lack of major health conditions or obstetrical complications in medical records.

### Pregnancy phenotypes

Medical records were used to determine birth outcomes. Gestational age in complete pregnancies were estimated using the American Congress of Gynecologists recommendations and previously described in greater detail [6,15,16]. The delivery less than 37 weeks gestation was defined to be preterm birth. Among preterm birth cases, we further disaggregated cases as spontaneous preterm birth cases if they had presentation of premature rupture of membranes, spontaneous preterm birth, or both.

### Sample preparation

Stored plasma samples, which were collected from the women between 24 and 28 weeks gestation and subsequently stored at -80C, were thawed on ice in preparation for analysis. Deproteinization was then performed by taking 100 µL of plasma combined with 400 µL 1:1:1 ratio of methanol, acetone, and water. Internal standards were also incorporated for metabolites recovery assessment, and included: 5 µM of L-(D4)Thymine, L-[^15^N] Anthranilic acid; and 20 µM of L-(^15^N)_2_ Tryptophan, Gibberellic acid, L-Epibrassinolide. Plasma samples were subsequently vortexed and centrifuged for 10 minutes at 15,000 x *g*. The supernatant of the centrifuged samples were transferred to a clean vial and dried using nitrogen gas. The dried samples were reconstituted to 50 µL.

### Liquid chromatography/mass spectrometry

All samples were randomly processed and assigned to an LC-MS queue using a computerized algorithm. The reversed phase (RPLC)-MS analysis was performed on an Agilent 1200 LC/6530 aTOF MS system (Agilent Technologies, Inc., Santa Clara, CA USA) with the Waters Acquity HSS T3 1.8 µ column (Waters Corporation, Milford, MA). Samples were analyzed twice, once in positive and negative ion modes. In positive ion mode runs, mobile phase A is 100% water that has 0.1% formic acid while mobile phase B is 100% methanol that has 0.1% formic acid. The formic acid is replaced with 0.1% (m/v) ammonium bicarbonate in negative ion mode runs.

### Metabolite identification

The “Find by Feature” algorithm is used to detect chromatographic peaks representative of metabolites by the Masshunter Qualitative Analysis Kit. Between samples, feature alignment was performed using an in-house written software package that matches features with identical mass and retention time between samples. In order to reduce gaps in chromatographic data, recursive feature identification was also performed by searching the data a second time with the list of aligned features using the “Find by Formula” algorithm in Agilent Masshunter Qualitative Analysis Software. Metabolites were putatively annotated using the mass spectral data annotation tool, Binner [17], to reduce contaminants, artifacts, and degeneracies. An annotated metabolite list was searched against an in-house library of 800 known metabolite standards which had been previously analyzed under identical LC-MS conditions. MS/MS spectra for metabolites not identified by standards were searched in the Metlin (Agilent Metlin B.08.00) or NIST 17. Metabolites not identified by library standards or MS/MS spectra were searched in the Metlin database (http://metlin.scripps.edu) and Human Metabolome Database (HMDB; http://www.hmdb.ca).

### Metabolomics data preprocessing

Samples were received in a single batch, with 333 metabolites species detected in total. Missing value imputation was performed using K-nearest neighbors (KNN) method [18]. Log-transformation and quantile normalization [19] were applied to the data, prior to the other downstream analysis,

### Source of variation analysis and data screen

The metabolomics dataset of maternal plasma consists of 333 metabolites. To eliminate confounders that are not truly related to preterm birth, we conducted a preliminary screen according to the source of variation (SOV) analysis, which helps to discover the contributions of each clinical/physiological factor to the metabolomics variation. The metabolites with a F statistic of preterm/control > 1 were screened before other analysis, meaning that they had a regression sum of squares larger than the error sum of square. All 333 metabolites passed this screening process.

### Differential metabolomics species identification

To remove potential confounding effects, we fit a linear model for each metabolite over preterm status while adjusting for *a priori* phenotypic variables via R *limma* package [20]. Adjusted phenotypic variables include gestational age in weeks, smoking status, alcohol consumption, baby length, baby gender, LGA/SGA (large/small baby for gestational age), maternal age, income, and pre-pregnancy BMI. As a result, 38 metabolites with p-values < 0.05 were selected as statistically significant in association with preterm birth.

### Weighted gene co-expression network analysis

For the weighted gene co-expression network analysis (WGCNA) analysis, all metabolites were analyzed together [21]. The smallest soft threshold with an adjusted R^2^ > 0.8 was 4, and hence it was chosen to calculate the adjacency score between any 4 metabolites within a sample set. Following that, the topological overlap value between these 4 metabolites was computed from this adjacency score and the corresponding connectivity value [22]. The topological overlap value is converted to a distance value by minusing it from 1 and producing a pairwise metabolites distance matrix. This distance matrix was then used to cluster the metabolites using hierarchical clustering with dendrogram, where modules were identified. As a result, we kept the metabolites with their topological overlap score larger than 0.5 in each module. For the integrated WGCNA analysis using both preterm and healthy samples, we used a soft threshold (power) of 8 as suggested by the WGCNA estimation. We setted minModuleSize 10, mergeCutHeight 0.25, deepSplit 2 and verbose 3 for the WGCNA analysis.

### The model of classification

We utilized the *Lilikoi* package [23] to determine the best machine learning model for classifying preterm and control samples using selected metabolites. Seven algorithms were compared in this step: recursive partitioning and regression trees (RPART), partition around medoids (PAM), gradient boosting (GBM), logistic regression with elastic net regularization (LOG), random forest (RF), support vector machine (SVM), and linear discriminant analysis (LDA). The samples were randomly split into 80/20 ratio for training data vs. testing data. The best method was determined on the training set using 10-fold cross-validation, by metrics F-statistics and balanced accuracy.

### The mapping of metabolite-related pathway and phenotype

We used the query lipid as the input to map metabolites to pathways from HMDB, PubChem, and KEGG in *Lilikoi [23,24]*. These metabolite-pathway interactions were then used for the further pathways analysis. Pathway dysregulation scores (PDS), a metric representing the degree of dysregulation at the pathway level, were calculated through the Pathifier R package to determine the dysregulation level of the pathway [25].

### Causality analysis

We sorted metabolomics data and clinical features into time series by the gestational ages of patients. Then we performed the Granger causality test to identify potential causality relationships between metabolites and preterm birth using the lmtest R package (version 0.9-37). The threshold of p-value is set to 0.01 for significant causality interaction.

## Results

### Study overview

The demographic and major clinical characteristics of the subjects in the PROTECT cohort study is shown in table 1. Except for the fact that individuals with preterm deliveries have significantly shorter gestational ages than healthy pregnant women (mean gestational age 39.20 weeks vs. 34.69 weeks, p-value = 1.28e-13), other characteristics of cases and controls are comparable across all categories. We also investigated the correlations among phenotypic factors (Fig. 1A). Income is positively correlated with preterm birth in weeks (PCC_Income_ = 0.205, p-value<0.05), confirming the social-economic association with preterm birth [26]. Maternal age shows the tendency of negative correlation with preterm (PCC_Age_ = -0.181, p<0.1).

**Table 1.**
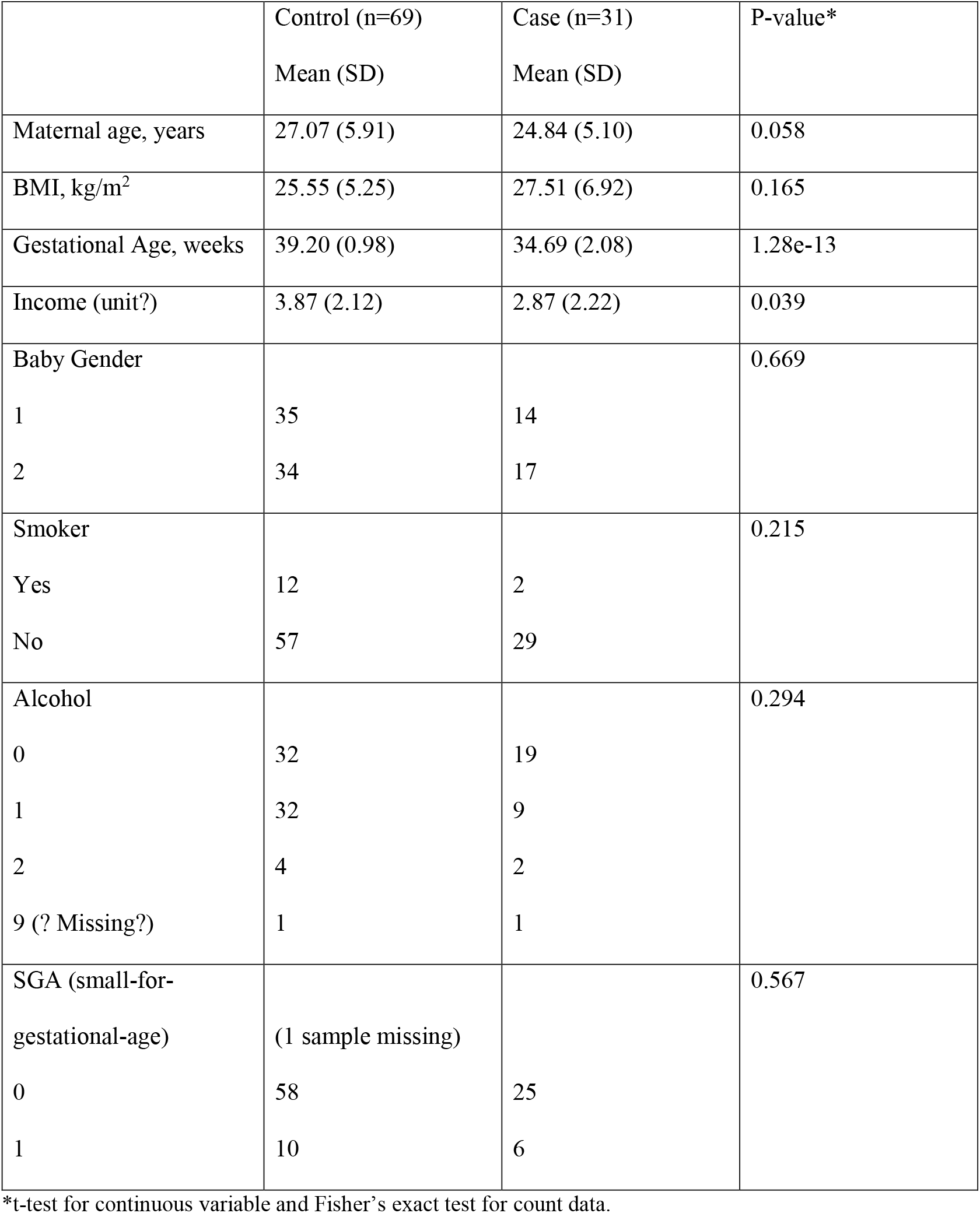
Demographic and clinical characteristics for subjects in this study (statistics are calculated after missing data imputation)

|  | Control (n=69)<br>Mean (SD) | Case (n=31)<br>Mean (SD) | P-value* |
| --- | --- | --- | --- |
| Maternal age, years | 27.07 (5.91) | 24.84 (5.10) | 0.058 |
| BMI, kg/m<sup>2</sup> | 25.55 (5.25) | 27.51 (6.92) | 0.165 |
| Gestational Age, weeks | 39.20 (0.98) | 34.69 (2.08) | 1.28e-13 |
| Income (unit?) | 3.87 (2.12) | 2.87 (2.22) | 0.039 |
| Baby Gender |  |  | 0.669 |
| 1 | 35 | 14 |  |
| 2 | 34 | 17 |  |
| Smoker |  |  | 0.215 |
| Yes | 12 | 2 |  |
| No | 57 | 29 |  |
| Alcohol |  |  | 0.294 |
| 0 | 32 | 19 |  |
| 1 | 32 | 9 |  |
| 2 | 4 | 2 |  |
| 9 (? Missing?) | 1 | 1 |  |
| SGA (small-for-gestational-age) | (1 sample missing) |  | 0.567 |
| 0 | 58 | 25 |  |
| 1 | 10 | 6 |  |

\*t-test for continuous variable and Fisher's exact test for count data.

**Figure 1.**
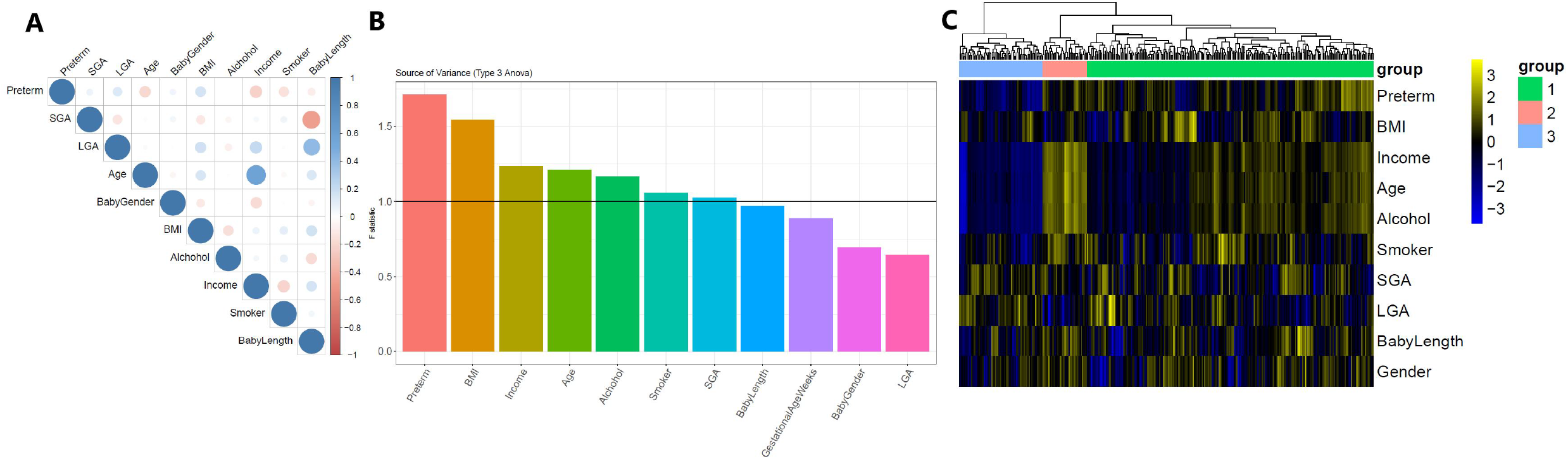
(A) Correlation matrix of the 10 phenotypic variables on the 100 samples (69 controls vs 31 preterm cases). (B) Source of variation (SOV) analysis using 100 samples. 333 metabolites are used in the ANOVA model. (C) Heatmap of correlations between 333 metabolites and 11 confounding factors. The rows represent the clinical factors, and the columns represent metabolites (Point-Biserial Correlation for continuous and binary covariates; Pearson Correlation for continuous covariates; Spearman Correlation for continuous and ordinal covariates).

A total of 333 metabolites were identified by LC/MS. To examine the degree of confounding from other variables, a source of variation (SOV) analysis was carried out (Fig. 1B). Preterm birth is ranked the first for the F statistics, followed by variables BMI, income, maternal age, alcohol consumption, smoking, and SGA which all have F statistics bigger than 1. To further identify the relationships between phenotypic factors and metabolites, correlations between clinical factors and metabolites were calculated (Fig. 1C) and then subject to hierarchical clustering (using Euclidean distance as the distance metric). Three Clusters of metabolites are identified with sizes of 231, 36, and 67. Cluster 3 is significantly enriched in fatty acids (FAs) (Fisher’s p-value = 5.24e-4, odds ratio = 2.12), and FAs are generally lower in preterm samples. They have a striking pattern of negative associations with preterm birth. Moreover, FAs also have overall negative associations with age, income, and alcohol use, suggesting the biological, social-economical, and behavioral effects are intertwined at the metabolomic level. The other two clusters do not have enrichment in specific metabolite functional groups.

### Correlation network analysis of metabolomics related to preterm birth

To further elucidate the relationships between metabolomics and preterm birth, we next performed the weighted gene correlation network analysis (WGCNA) method on the 333 metabolites [21]. WGCNA analysis yields 7 modules (Fig. 2A). Among these modules, only the turquoise-colored module shows a significant positive association (Fisher’s Exact Test, p-value=0.08e-02) with preterm birth (Fig. 2A-B). This module is enriched with FAs (Fisher’s Exact Test, p-value=3.85e-05) and carene (CAR) (Fisher’s Exact Test, p-value=2.53e-03). This FA/CAR enriched module also shows a significant negative association (p-value=0.002) with gestational age (GestAge) (Fig. 2B). These results, together with the previous metabolite-phenotype analysis (Fig. 1C), demonstrate that FAs in the mothers who gave birth prematurely not only have higher levels but also tighter correlations (through regulations). To examine the module difference between cases and controls more closely, we further conducted the WGCNA on the two groups separately. Three modules have significantly overlapping metabolites in the case and control groups (FigS1. 2A and 2B), respectively. Interestingly, the FA enriched modules in cases (A2) and controls (B2) have the most significant overlap (p-value=6.76e-18) (FigS1. 2C). However, we did not find that the density of FA-enriched modules was higher in preterm cases compared to that in control (FigS1. 2D).

**Figure 2.**
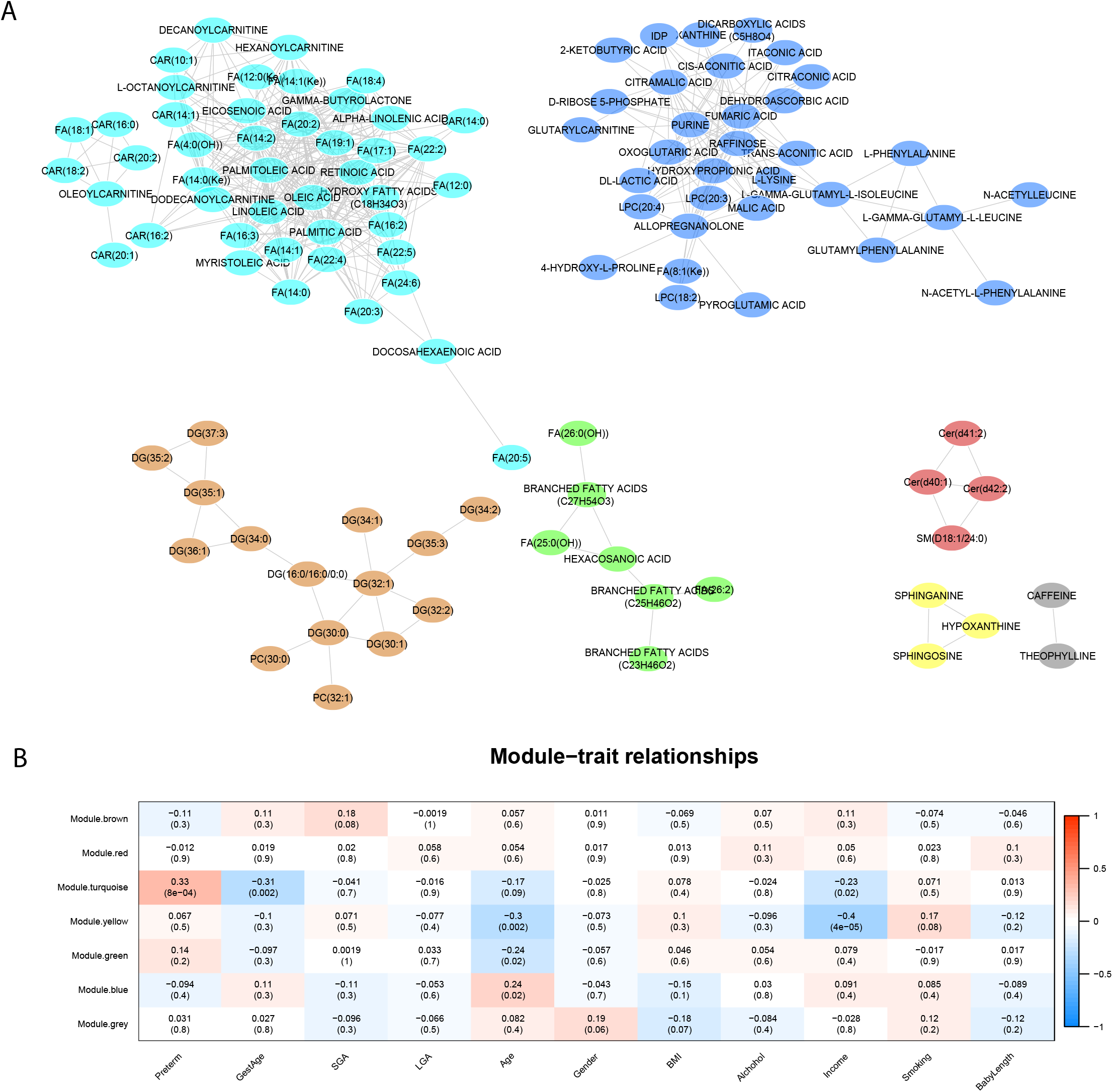
WGCNA network in all samples. (A) WGCNA network modules of metabolomics data from both preterm and control samples. Each node represents a lipid. Node color represents a module. (B) Module-trait associations.

### Differentiated metabolites and their mapped pathways

We next conducted differential metabolite analysis between cases and controls, using *limma* package [20] allowing for phenotypic variable adjustment. As a result, 38 metabolites are significantly different (p-value <0.05) between preterm and control samples exclusively, and are not associated with other confounders (Fig. 3A). Among them, 21 metabolites are up-regulated and 17 metabolites are down-regulated in preterm samples (Fig. 3B). The majority of these metabolites are unsaturated fatty acids.

**Figure 3.**
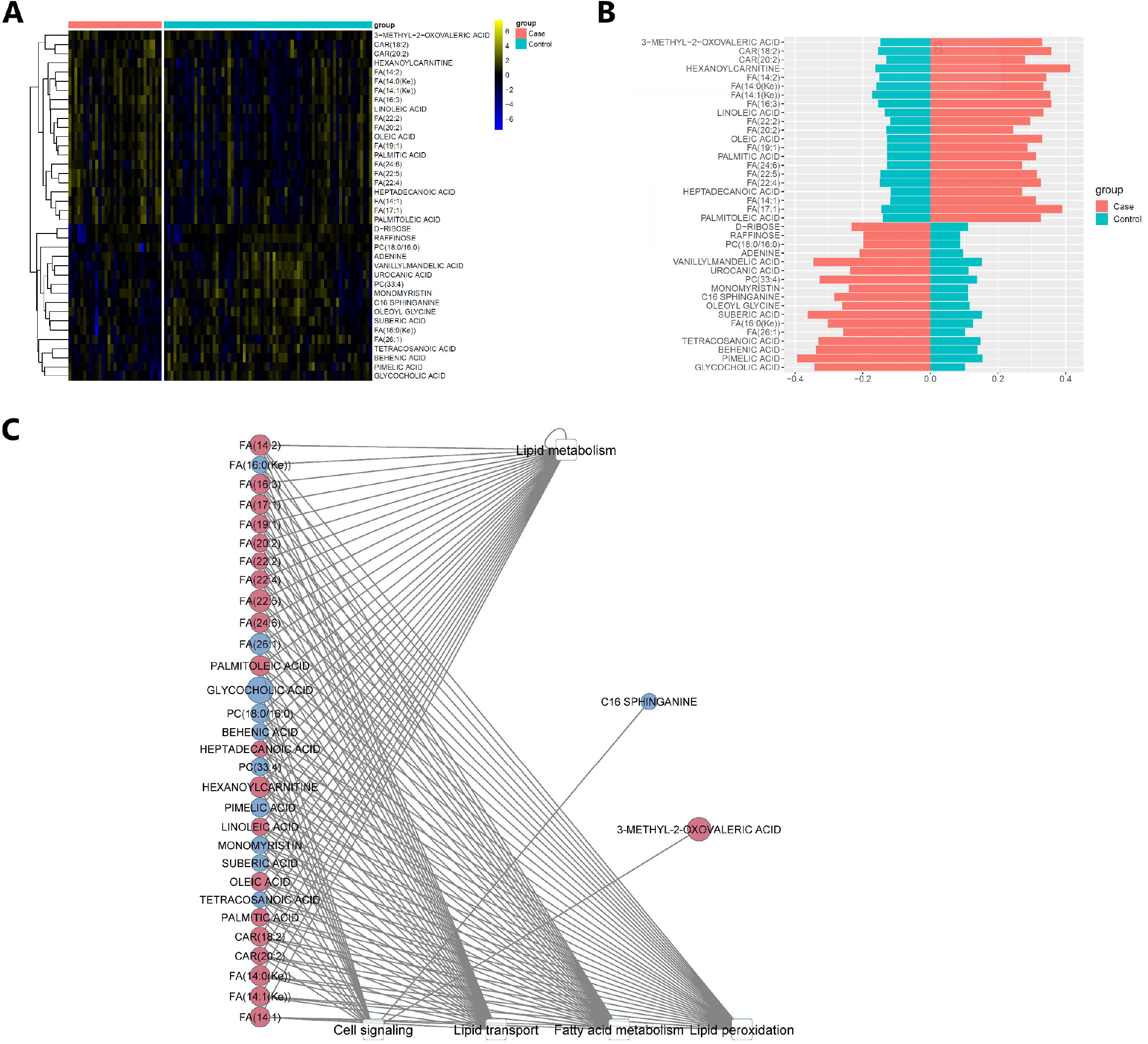
Metabolites show significantly different levels in preterm and control samples. (A) Heatmap of the 38 metabolites with a significant difference exclusively between preeclampsia and control samples (p-value <0.05). (B) Barplots on the averaged normalized intensities in cases vs controls. (C) Bipartite graph of the significantly differentiated metabolites and the significantly altered metabolic pathways they are associated with. Five pathways with a significant difference between preterm and control samples (p-value <0.05) and 33 significantly differentiated metabolites engaged in these pathways are shown. Elliptical nodes: metabolites. Rectangular nodes: pathways from HMDB, PubChem, and KEGG databases. Node color: Red, up-regulated; Blue, Down-regulated. Node size: the absolute value of log fold change (logFC).

To further explore the functions of these metabolites, we mapped the 333 metabolites to pathways and conducted pathway enrichment analysis, using the *Lilikoi* R package [18,23]. These pathways are from KEGG, HMDB, metlin and pubchem databases. 240 out of 333 metabolites are successfully mapped by at least one database, with assigned memberships to 38 pathways. Among the 38 differential metabolites, 33 of them are involved in 5 pathways that show significant alterations in pathway dysregulation scores, a metric representing the degree of dysregulation at the pathway level [25]. These pathways share a lot of lipids and are interrelated: Lipid metabolism, cell signaling, lipid transport, fatty acid metabolism and lipid peroxidation. The bipartite plot illustrated the relationships between the differentiated metabolites and their corresponding differential pathways (Fig. 3C).

### Metabolomics based preterm biomarker model

Another important application of metabolomics analysis is to screen for diagnostic biomarkers for diseases. For this purpose, we split samples with 80/20 ratio into training and testing data. We further selected 17 metabolites out of the 38 differentiated ones using mutual information score 0.5 as the threshold. We compared the performance of seven machine learning algorithms in the *Lilikoi* R package, including recursive partitioning and regression trees (RPART), partition around medoids (PAM), gradient boosting (GBM), logistic regression with elastic net regularization (LOG), random forest (RF), support vector machine (SVM), and linear discriminant analysis (LDA). We used the area under the ROC curve (AUC), F1 statistic and balanced accuracy to evaluate the models. Among all classification methods, RF yields the highest balanced accuracy statistic (1.0) in the training dataset (Fig. 4A), so we selected it as the winning model to show the predictive performance on the remaining testing dataset. The overall accuracy for RF on the testing data is 0.92 for the AUC, 0.5 for the F1 statistic, and 0.67 for the balanced accuracy (Fig. 4C). Next, we tested if the biomarkers are specific to preterm birth rather than other clinical confounders. We used the 17-feature RF classification model built for preterm birth to predict its classification performance over other terms including LGA, BMI and maternal age, using the same testing data set. The AUC on LGA, BMI, and maternal age are 0.2, 0.09, and 0.17 respectively in the precision-recall curves (Fig 4D). This confirms the specificity of the 17-biomarker model for preterm birth. Several fatty acids show top importance scores in the model: FA(17:1) (1^st^, importance score = 7.32 out of 100); FA(24:6) (2^nd^, 7.02); FA14:2 (3^rd^, 6.98). Hexanoylcarnitine is also a top important metabolite (5^th^, 6.6), involved in fatty acid oxidation. It has been reported to be significantly higher in preterm birth [27].

**Figure 4.**
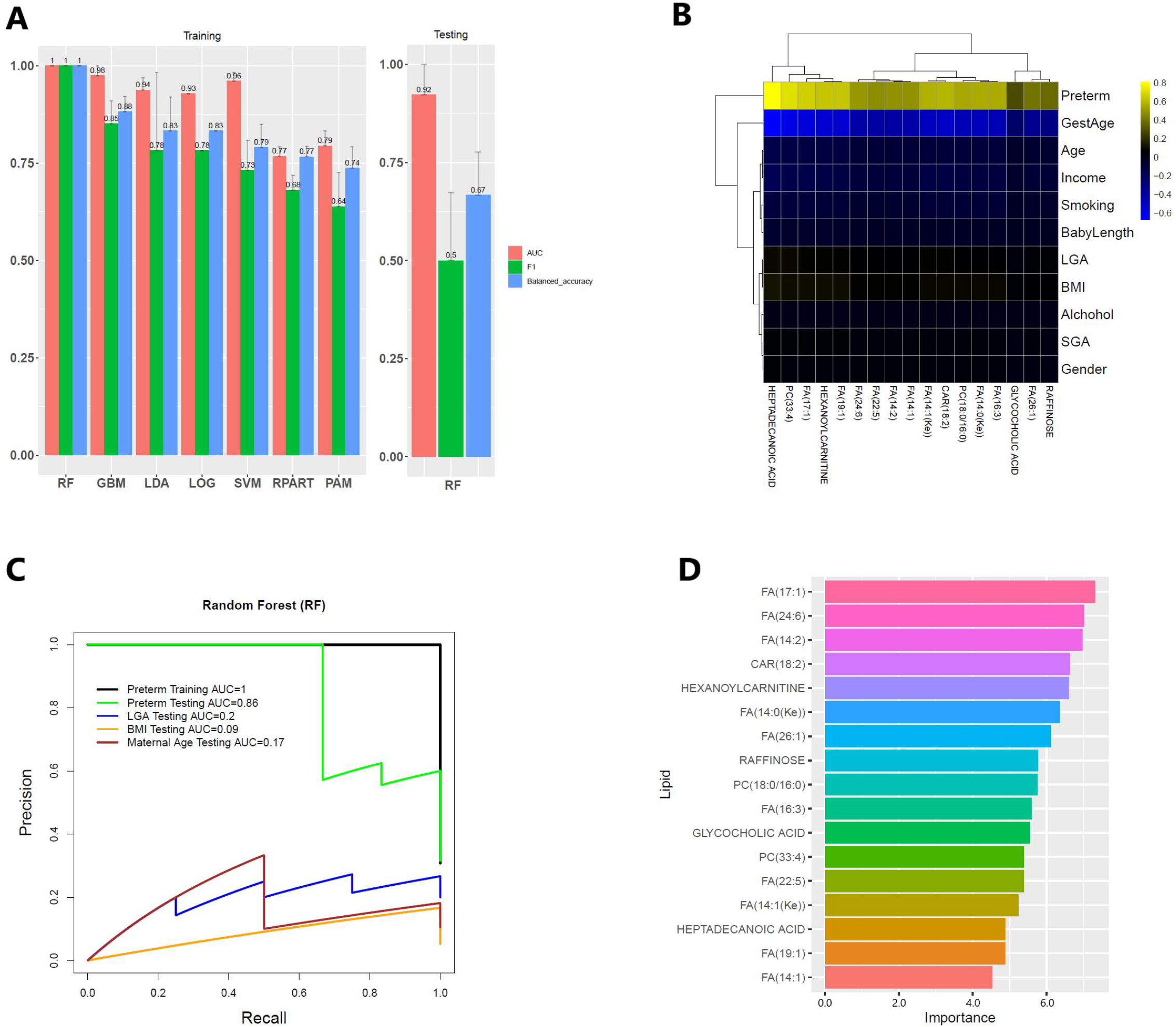
Classification model for preterm birth. (A) Comparison of seven classification models using 17 metabolites on the hold-out testing. The data set was randomly splitted into training data (80%) and testing data (20%) for 10 times. The average value and standard error of the 10 repeats are shown for three performance metrics of area under the ROC curve (AUC), F1 statistic, and balanced accuracy. The winning method RF in training data (left) was then applied to the testing data (right). (B) The heatmap of correlation coefficients between the 17 metabolites and clinical variables. (C) The precision-recall curves of the RF model from (A) on classifying preterm, LGA (large for gestational age), Income and Maternal Age (>=35 yrs or not) respectively, using the same set of testing data as in (A). (D) Normalized variable importance scores for the 17 lipid markers in the RF model. The normalization is done on R by making the sum of importance scores to be 100.

### Predicted causality interactions among metabolites and preterm birth

We used the Granger causality test [28] to infer significant causality interactions (p-value < 0.01) between the 17 metabolites and preterm. As shown in Fig. 5, up-regulated hexanoylcarnitine, CAR(18:2), CAR(20:2), FA(14:1(Ke)), FA(14:2), FA(17:1), and down-regulated behenic acid, pimelic acid, suberic acid, glycocholic acid, and PC(33:4) are predicted as direct casual metabolites of preterm birth. The causality test also predicts the causality interaction from FA(17:1) to pimelic acid, which is synthesized from fatty acid [29]. Interestingly, down-regulated suberic acid is predicted to be the direct cause of up-regulated FA(22:4), FA(20:2), FA(22:2), FA(14:0(Ke)), FA(14:1(Ke)), and FA(14:2). A previous study shows that suberic acid is present in the urine of patients with fatty acid oxidation disorders, indicating the correlation between suberic acid and the metabolism of fatty acids [30].

**Figure 5.**
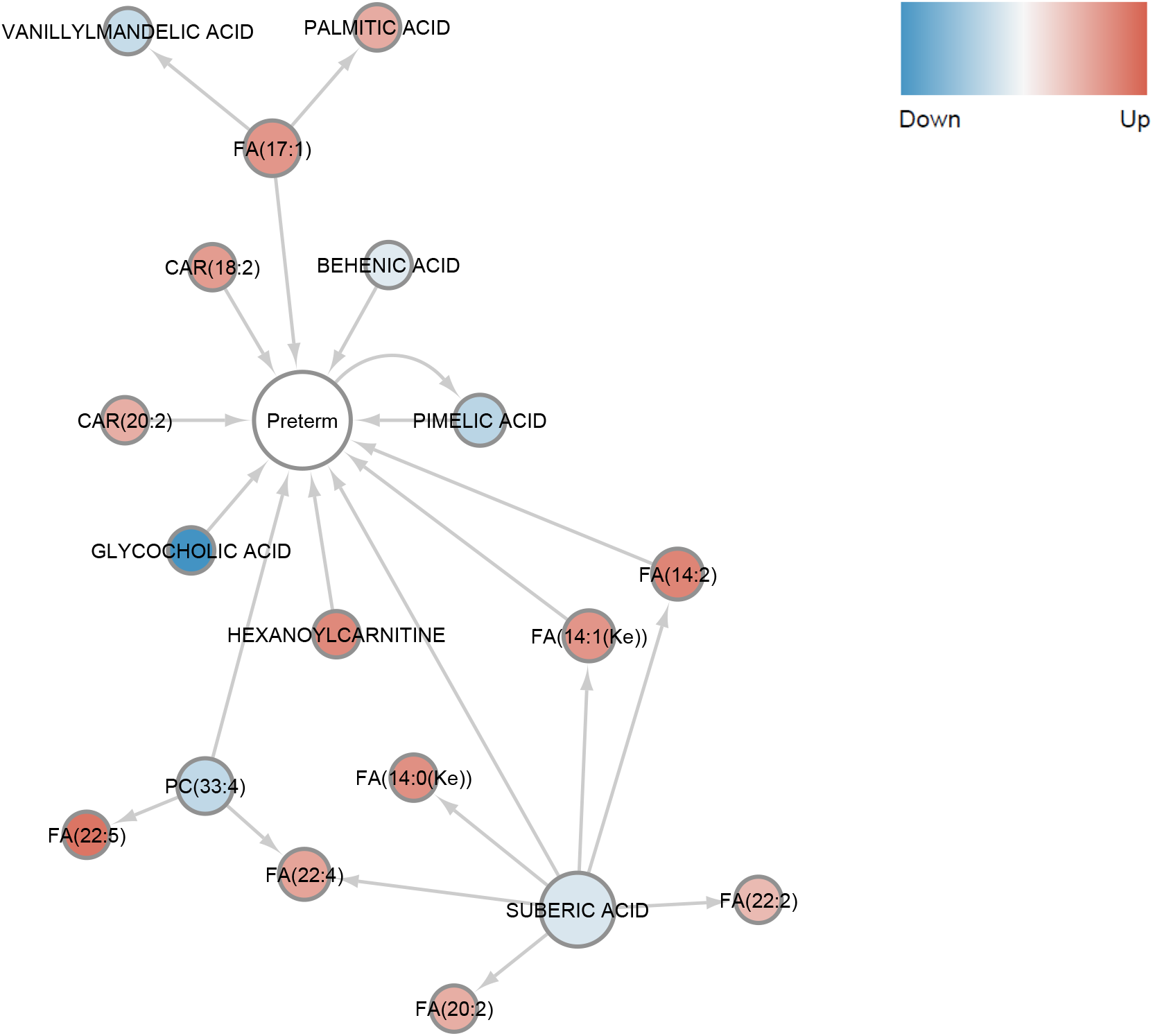
Predicted significant (p-value < 0.01) causality interactions between the 17 metabolites and preterm birth. Arrow indicates the causality interaction. Blue and red nodes are down and up-regulated metabolites, while the center one is preterm.

## Discussion

Preterm birth is one of the leading causes of newborn mortality and morbidity [1]. To improve our understanding of preterm birth, we conducted metabolomics analysis of maternal blood in the PROTECT cohort of preterm birth patients and healthy controls.

The importance of fatty acids in preterm birth is highlighted by bioinformatics analysis in various aspects. First, correlation network analysis of metabolomics reveals deregulated lipid modules that may contribute to preterm birth (Fig. 2). The FA/CAR enriched module is enriched with several fatty acids including two essential fatty acids, i.e. alpha-linolenic acid and linoleic acid (omega-6 fatty acid), and a class of saturated fatty acids (SFAs: heptadecanoic acid, palmitic acid). Other studies have also found excessive free fatty acids detected in the preterm cases of maternal circulation, linking them to inflammation [31] the main cause of preterm birth [27]. In fact, a higher omega-6 to omega-3 fatty acid ratio would increase pro-inflammatory eicosanoid production [32,33] and it was associated with shorter gestation duration for overweight/obese women [34]. Another study on underweight and obese women with spontaneous preterm birth identified a higher concentration of omega-6 and omega-3 fatty acids in their mid-gestation serum samples [35]. Confirming our discovery, a recent complementary lipidomics study within the PROTECT cohort also observed that mono- and poly-unsaturated free fatty acids (FFA 20:1, FFA 20:1, FFA 18:1) were associated with a higher risk of spontaneous preterm birth [14]. We have also found complementary evidence in the LFIECODES cohort of positive associations between spontaneous preterm birth and eicosanoids, which are secondary metabolites of poly-unsaturated fatty acid parent compounds such as arachidonic acid [36]. Besides fatty acids, two phosphatidylcholine (PC(18:0/16:0), PC(33:4)) were also selected by the biomarker model for preterm birth. These two metabolites have lower levels in preterm births. PCs are the main structure of cell membranes and play an important role in maintaining membrane stability and reducing inflammation [37]. Consistent with this, one recent study also found a class of PC significantly lower in preterm births [38].

Interestingly, the causality analysis shows the causal effect of decreased suberic acid for the excessive fatty acids. This is consistent with a previous finding that suberic acid is related to fatty acid disorders [30]. Suberic acid, also called octanedioic acid, is a dicarboxylic acid, which can be produced from fatty acids [39]. The production from fatty acids to dicarboxylic acids are catalyzed by cytochrome P450 (CYP) 4 F/A (CYP4F/A) enzymes [39,40]. The accumulation of fatty acids and reduction of suberic acid in preterm maternal blood samples (Fig 5) suggest that CYP4F/A enzymes, the enzyme catalyzing this conversion, have reduced activities in preterm delivery. Polymorphisms in CYP4F/A genes, which impair enzyme functions, previously showed associations with preterm birth [41]. Thus, we speculate that polymorphisms or other forms of deactivation of CYP4F/A genes may play a role in preterm births.

Changes in these lipids collectively suggest that lipid metabolism may contribute to the pathogenesis of preterm birth (Fig 6). Indeed, several related pathways including lipid metabolism, fatty acid metabolism, and lipid peroxidation pathways are all enriched in the preterm cases (Fig 3C). These pathways were discussed frequently in many previous preterm birth analyses [38,42,43]. UFAs, shown to be excessive in preterm samples of this dataset, are more likely to undergo lipid peroxidation[44]. UFAs and the evident lipid peroxidation process could lead to oxidative stress, which was reportedly related to preterm birth through regulating ripening cervical, uterus contraction, and membrane rupture [42]. and accelerated lipid peroxidation is found in prematurity [45].

**Figure 6.**
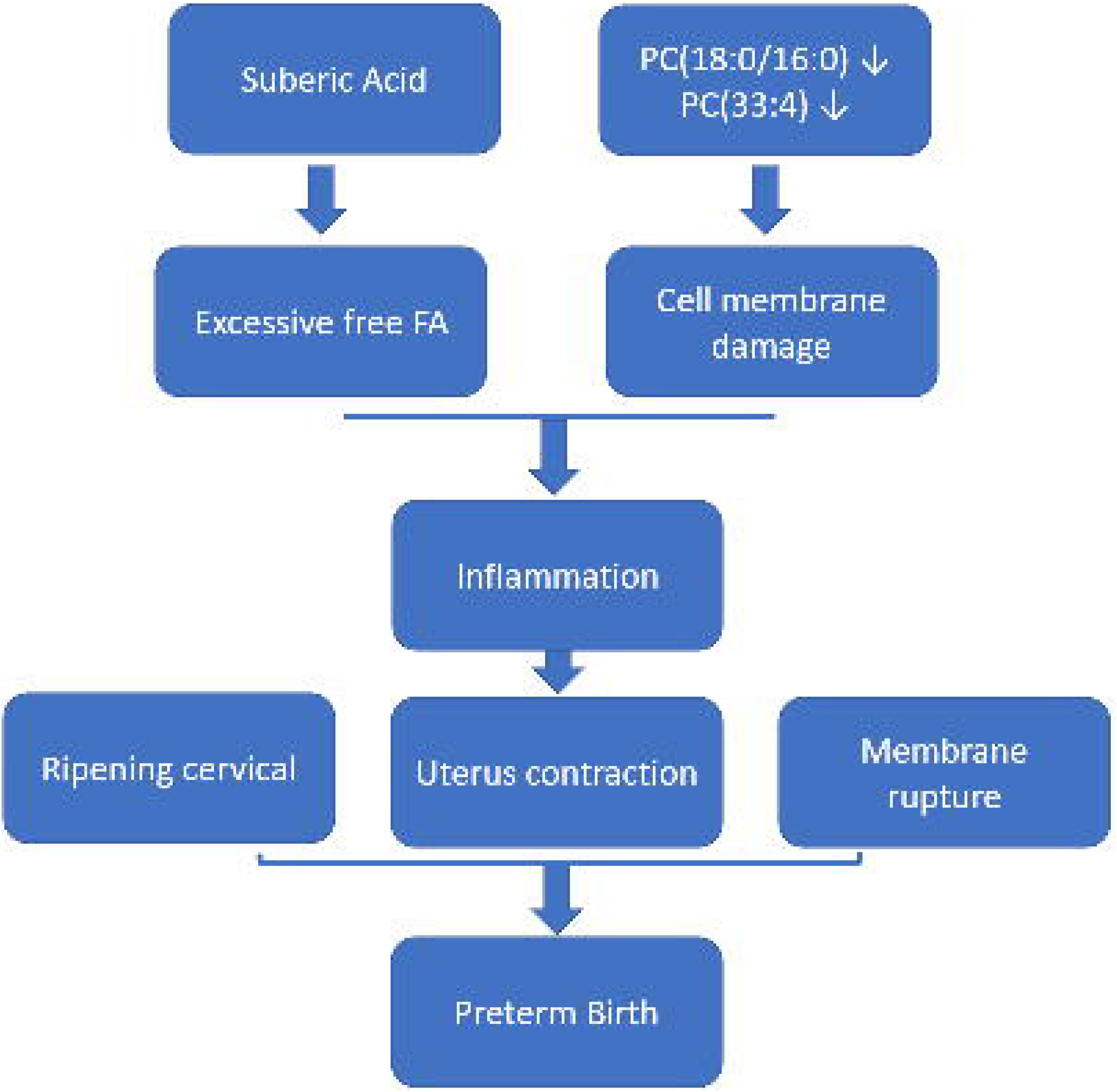
A proposed model of metabolites changes affecting preterm birth.

A few caveats of this study should also be mentioned. First, subjects in this study do not have specific dietary records, thus potential confounding from diets cannot be investigated. The metabolites are measured from maternal blood, therefore any biological mechanisms discussed here are inferred systematically rather than being directly measurable from relevant tissues (eg. placenta). Additionally, despite developing a potential biomarker panel from a classification model, these candidates are suggestive and not quantitatively validated yet. We plan to validate them in other independent cohorts in the future. Nonetheless, this study provides strong evidence of the involvement of a class of saturated and unsaturated FAs and PCs in preterm births, mediated by perturbation in biological functions including cell signaling and lipid peroxidation.

## Data Availability

The metabolomics data set has been uploaded to Metabolomic Workbench, which is a public repository for metabolomics.

http://dx.doi.org/10.21228/M8DH5P.

## Availability of source code and requirements

Project name: Maternal lipids in the pathogenesis of preterm birth

Project home page: https://github.com/lanagarmire/pretermBirth_metabolomics

Operating system(s): Windows and Linux

Programming language: R

## Data Availability

The metabolomics data set has been uploaded to Metabolomic Workbench, which is a public repository for metabolomics, at http://dx.doi.org/10.21228/M8DH5P.

## Abbreviations

LDA: linear discriminant analysis
RF: random forest
LOG: elastic net
GBM: gradient boosting
SVM: support vector machine
RPART: classification tree
PC: phosphocholine
PS: acylglycerophosphoserines
PE: diacylglycerophosphoethanolamines
PI: phosphatidyinositol
PG: phosphatiduglecerol
FA: fatty acid
CAR: carene
CYP4F/A: cytochrome P450 (CYP) 4 F/A
AUC: area under the ROC curve
WGCNA: weighted gene correlation network analysis
SOV: source of variation

## Competing Interests

The authors declare that they have no competing interests.

## Author’s contribution

YC and BH conducted the bioinformatics analysis, modified code provided by YL. MA provided writing material. JDM designed the study, obtained funding, supervised the metabolomics assays, and critically reviewed early drafts of the paper. LXG supervised the analysis. YC, BH and LXG wrote the manuscript. All authors have read and revised the manuscript.

## Funding

This study was supported by the Superfund Research Program of the National Institute of Environmental Health Sciences, National Institutes of Health (grants P42ES017198). Additional support was provided from NIEHS grant numbers P50ES026049, R01ES032203, and P30ES017885 and the Environmental influences on Child Health Outcomes (ECHO) program grant number UH3OD023251. LXG is supported by grants K01ES025434 awarded by NIEHS through funds provided by the trans-NIH Big Data to Knowledge (BD2K) initiative (www.bd2k.nih.gov), R01 LM012373 and LM012907 awarded by NLM, R01 HD084633 (LXG and SS) awarded by NICHD.

## Acknowledgment

We thank the nurses and research staff who participated in cohort recruitment and follow up, as well as the Federally Qualified Health Centers (FQHC) and clinics in Puerto Rico who facilitated participant recruitment, including Morovis Community Health Center (FQHC), Prymed: Ciales Community Health Center (FQHC), Camuy Health Services, Inc. (FQHC), and the Delta OBGyn(Prenatal Clinic). We thank the BRCF Metabolomics core of University of Michigan for providing the assays and helping upload the metabolomics data to the public repository.

## Supplementary materials

Supplementary Figure S1. (A-B) WGCNA network in preterm births (A) and healthy controls (B), respectively. Each node represents a metabolite, whose size is proportional to the node connectivity value in a WGCNA network. (C) The overlap between modules of networks in control and preterm samples. (D) Detailed information on overlapping module density discovered in (C). (E) Bar plot of the connectivity scores of the 17 up-regulated metabolites.

## Notes

### Competing Interest Statement

The authors have declared no competing interest.

### Author Declarations

Ethics and Research Committees of the University of Puerto Rico (IRB # A8570110) and Northeastern University (IRB # 150629).

