## Supplementary figures and images for "Maternal plasma lipids are involved in the pathogenesis of preterm birth"

### Fig S1

Fig2 A

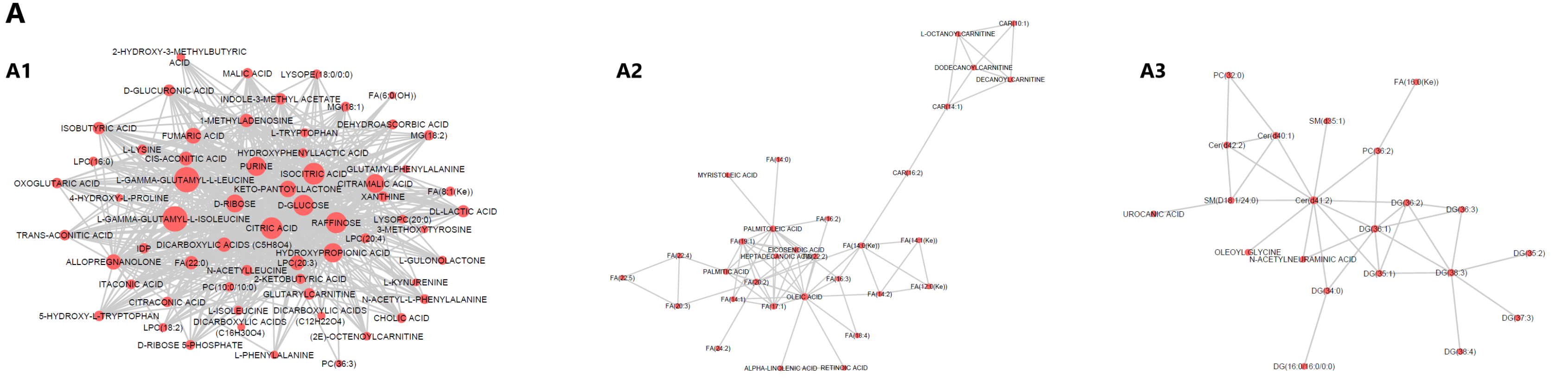


B

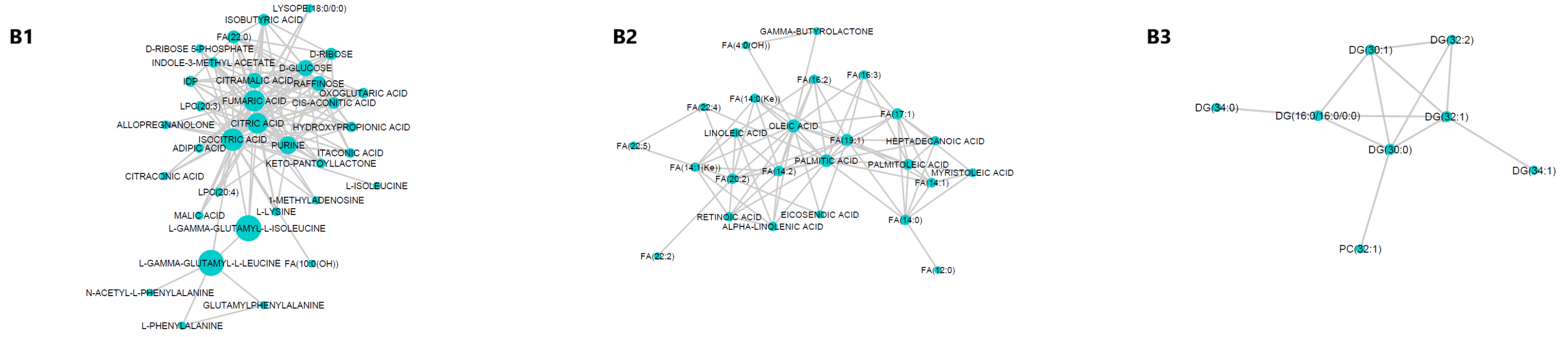


C

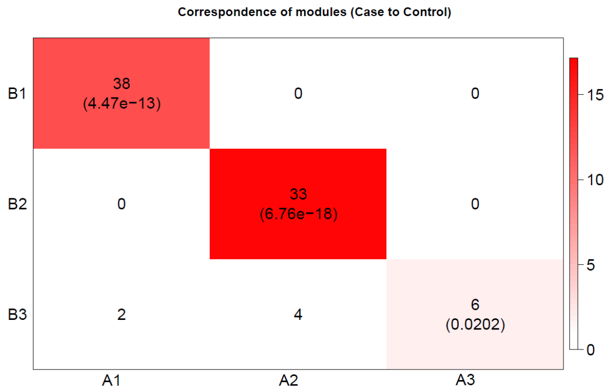


D

| | | Preterm | Controls |
|---|---|---|---|
| Module Density | Module 1 (38) | 0.266 | 0.169 |
| | Module 2 (33) | 0.078 | 0.138 |

E

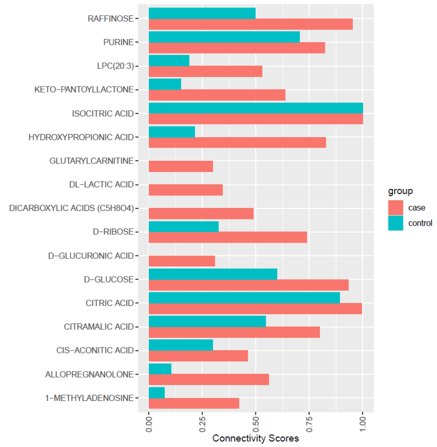
